# Water Insecurity, Water Borrowing, and Psychosocial Stress Among Daasanach Pastoralists in Northern Kenya

**DOI:** 10.1101/2022.01.26.22269937

**Authors:** Leslie Ford, Hilary J Bethancourt, Zane Swanson, Rosemary Nzunza, Amber Wutich, Alexandra Brewis, Sera Young, David Almeida, Matthew Douglass, Emmanuel K. Ndiema, David R. Braun, Herman Pontzer, Asher Y. Rosinger

## Abstract

This article quantifies Daasanach water insecurity experiences in Northern Kenya, examines how water insecurity is associated with water borrowing and psychosocial stress, and evaluates if water borrowing mitigates the stress from water insecurity. Of 133 households interviewed in 7 communities, 95% were water insecure and 74.4% borrowed water three or more times in the prior month. Regression analyses demonstrate water borrowing frequency moderates the relationship between water insecurity and psychosocial stress. Only those who rarely or never borrowed water reported greater stress with higher water insecurity. The coping mechanism of water borrowing may help blunt water insecurity-related stress.

## Introduction

Between 1990 and 2015 the United Nations estimates that 2.6 billion people gained access to improved drinking water sources globally (United Nations, n.d.). Despite this, 4 billion people still lack sufficient safe water for at least one month per year (Mekonnen & Hoekstra, 2016), and 2.2 billion people obtain their water from an unsafe source (UNICEF and WHO, 2019). With global water need expected to increase at a rate of approximately 1% annually over the next 30 years (UN Water, 2019), a growing number of people may have to rely on non-institutional or informal systems to meet their water needs (Rosinger et al. 2020).

Shortages of clean safe water are the result of both natural water distributions, such as climate shifts and geography, and social influences, including political structures, cultural norms, and technical interventions (Zwarteveen et al., 2017). Climate shifts and extreme weather events have led to some areas becoming hotter and drier while others are experiencing catastrophic flooding (UN Water, n.d.; Konapala et al, 2020). This is compounded by changing political structures, which influence the ways in which water is distributed, favoring some and further marginalizing others, particularly in rural regions (Wutich et al., 2017). Cultural norms around gender and water governance (Brewis et al., 2019a), as well as implementation of exploitive technologies such as deep wells or dams, have also shaped water distributions and availability.

Insufficient access to clean water has been linked to proximate acute health conditions (Rosinger & Young, 2020), such as dehydration and diarrhea (Rosinger, 2018), chronic conditions, including hypertension (Brewis et al., 2019a; Rosinger et al., 2021), and psychosocial and emotional distress (Collins et al., 2019; Boateng et al., 2020; Stevenson et al., 2012; Wutich, 2009). Both acute and chronic health outcomes linked with water insecurity have historically affected those in the global south disproportionately (Wutich et al., 2017). Thus, understanding how populations cope with water insecurity is critical for untangling the relationship between water insecurity and psychosocial stress.

### Water insecurity and Coping Strategies

Water insecurity, or the “inability to access or gain adequate, reliable, and safe water for wellbeing and a healthy life” (Jepson et al., 2017), varies by context. Operationalizing this concept relies on the societal discourse and cultural norms surrounding water and water procuring practices in each context (Pahl-Wostl, Gupta, & Bhaduriwater, 2016). In many areas, water insecurity experiences are poorly documented or understood (Wutich et al., 2017). These experiences may relate to individual and collective decisions about the acquisition, distribution, and prioritization of water for different water needs and users (Collins et al., 2019).

Water insecurity coping strategies, or actions taken by a household in response to unreliable water supplies (Majuru et al. 2016), are strategies used to reduce water problems and improve access (Venkataramanan et al., 2020). In a recent meta-ethnographic synthesis of relevant qualitative literature, Achore and colleagues (2020), identified nine common coping strategies for water insecurity. The type of coping strategies employed varied by income. Wealthier households often choose “hard” coping strategies, which are more costly or technical solutions, such as water storage, construction of alternative water sources, buying water from private vendors, illegal connections to public water networks, rainwater harvesting, and water treatment to improve the quality. Poorer households often choose “soft” or behavioral interventions, which are less expensive but more time consuming. These coping strategies include water borrowing from social networks, water management and reuse, and fetching water from distant sources (Achore et al., 2020).

While necessary for meeting their immediate water needs, many of these coping strategies have tradeoffs that affect income-generating or leisure potential as well as having implications for water-insecurity related stress. The cumulative experience of water insecurity has been linked to emotional distress and poor mental health outcomes (Wutich & Ragsdale, 2008; Wutich et al., 2020). Due to their potential psychosocial toll, many water coping strategies are not sustainable as permanent solutions for water insecurity (Achore et al., 2020; Venkataramanan et al., 2020). For example, many household-level water managers experience psychosocial stress as a byproduct of the negotiations in which they must engage to ensure sufficient water supplies in the household (Wutich & Ragsdale, 2008). This link between psychological distress and water insecurity has been documented around the world including in Cochabamba, Bolivia (Wutich & Ragsdale, 2008), Lake Urmia, Iran (Zenko and Menga, 2019), Nepal (Brewis et al., 2019a) and Nyanza, Kenya (Collins et al., 2019). Thus, the coping strategies used, like water borrowing, may contribute to how individuals experience water insecurity, including the level of psychological burden (Brewis et al., 2019a; Stevenson et al., 2016; Stoler et al., 2019).

### Water Sharing and Water-Related Stress

Water sharing is an often “invisible” coping strategy used to meet water needs (Wutich et al., 2018; Rosinger et al., 2020). Water borrowing networks emerged from the traditional adaptation of pooling resources among vulnerable, often marginalized communities to meet collective water needs (Wutich et al., 2018). As a result, these communities often self-organize their water governance systems, which ensure a more equitable distribution of their limited water resources (Brewis et al., 2019b).

Even the small amounts of water that get transferred through household-to-household water sharing can have health implications. Sharing water between households has been linked to decreased dependence on inadequate water quality and reduced psychosocial stress (Wutich et al., 2018). It has also been linked with increased burden and distress during times of need through the creation of differentiated networks that include some and exclude others (Wutich et al., 2018; Stoler et al., 2019; Brewis et al., 2021). It is often through these networks that social groups use natural resources, such as water, to reinforce political, economic, and social barriers between those with authority and those without (Rademacher, 2015). Historical systems, cultural norms, ethnicity, and socio-economic status are just a few of the factors that (re)produce and reinforce power relations and subsequently determine who has and who does not have access to water (Cole, 2017).

Previous research has demonstrated that water borrowing acts as a coping strategy for water insecurity globally in communities with water problems (Rosinger et al. 2020). Pearson and colleagues (2015), found that among pastoralists in southern Uganda water sharing and reciprocity were crucial for meeting water needs. However, it remains unclear if engaging in water borrowing helps alleviate psychosocial stress from water insecurity. Few studies have examined this problem in pastoralist communities who often have conflicts with neighboring groups over water resources (Pearson et al., 2015; Straight et. al., 2016, Balfour et al., 2020). *Pastoralists and Water Insecurity*

Eastern African pastoralists have lived in semi-arid environments for millennia (Hildebrand et al., 2018). Among pastoralist communities, climate resilience has historically been linked to their mobility and broad social networks (Robinson & Berkes, 2010; Pearson et al., 2015). These adaptations allowed them to mitigate the impact of water scarcity by moving herds, limiting herd size, or selling livestock for goods needed to survive during periods of limited water availability (Wright, 2019). Yet, with changing environmental and social conditions, many pastoralists may no longer be able to rely on previous strategies, such as moving their homes and livestock, to cope with water shortages (Wright, 2019). Instead, some pastoralist groups have had to resort to alternative solutions, such as buying, sharing, or changing to distant but more reliable water sources in order to cope with water shortages (Pearson et al., 2015; Wutich et al., 2018). However, we do not know how these strategies may increase or reduce their psychosocial stress associated with water insecurity.

Therefore, this paper aims to fill these gaps by examining the water insecurity experiences, water sharing practices, and psychosocial stress of Daasanach pastoralists in Northern Kenya. We focus on Daasanach communities because they are an underserved population who have a long history of living in a water-scarce environment with limited resources (Kiura, 2008). We examine a critical “soft” coping strategy, water borrowing, to see if and how it is used to mitigate water insecurity-related stress.

We first describe the water insecurity experiences of Daasanch and how environmental and household characteristics are associated with their water insecurity scores. Second, we examine how water insecurity is associated with frequency of water borrowing. Finally, we examine perceived stress, and explore if water borrowing is an effective coping mechanism to decrease water insecurity. We test whether frequency of water borrowing moderates the relationship between water insecurity on psychosocial stress. We hypothesized that greater degrees of household water insecurity will be associated with higher psychosocial stress (Wutich & Ragsdale, 2008; Zenko & Menga, 2019; Brewis et al., 2019a; Boateng et al., 2020), but that higher frequency of water borrowing levels will buffer this relationship (Stoler et al., 2019).

## Materials and Methods

### Study Population

Daasanach are semi-nomadic pastoralists who have begun to adapt a semi-sedentary lifestyle. Their primary livelihood is herding livestock including goats, cattle, and camels (Sagawa, 2010). Daasanach communities inhabit the North-Eastern shores of Lake Turkana in present-day northern Kenya and southern Ethiopia. The Lake Turkana basin is home to several distinct pastoralist groups that share common grazing territories and water sources for livestock. These pastoralist groups have a long history of mitigating climate changes and water shortages through migrating with livestock to find alternative water sources and pasture as needed (Kefale & Gebresenbet, 2012; Hildebrand et al., 2018). Based on archeological records, pastoralists in the Turkana basin have been highly resilient to changing environmental conditions; however, that resilience has been undermined in the last century by changing social and climatic conditions (Wright, 2019).

Further, migration has become increasingly difficult due to external political, social, economic, and environmental influences (Sagawa, 2010). For example, the construction of the Gibe III hydropower dam in Ethiopia and the demarcation of the Sibiloi National Park in Kenya have created challenges for Daasanach. The Gibe III dam was constructed on the Omo River, which is the primary water source for Lake Turkana. This construction has dramatically lowered the water table, contributing to reduced vegetation for livestock in the Lower Omo Valley (Mwamidi et al., 2018) and increased the salinity of Lake Turkana. Non-governmental organizations have been attempting to increase water access for Daasanach with construction of bore-holes and wells in and around their main settlement as well as in the fora (Rosinger et al., 2021). The demarcation of their land for conservation has also curtailed mobility patterns hence limiting access to water resources (Greiner 2012). As a result, Daasanach and other tribes have been experiencing water shortages and increased conflict over limited resources (Hodbod et al., 2019). Recent work has demonstrated that Daasanach experience high levels of food and water insecurity, which indicates that they may be experiencing high levels of perceived stress as well (Bethancourt et al., 2021).

### Design and Sample

Interview and survey data were collected in June and July 2019 with follow-up community discussions and observations in October, 2020 and May, 2021. The months of June– August are after the long rainy season, making it an ideal time to observe and collect data on water insecurity. For this study, six permanent Daasanach communities and one temporary camp were selected as sites out of roughly 26 communities from which to recruit participants. These seven communities are located at varying distances from the town of Illeret, located on the shore of Lake Turkana and is the largest settlement in the area.

With the assistance of community partners including local elders and community health assistants or volunteers, we randomly selected every third household in each community, and extended an invitation to participate in this study. If the first household sampled was not home or declined to participate, the next household was invited. Between 12 and 28 households were sampled in each community depending on community size and amount of time spent in each location. Full study design details are described elsewhere (Bethancourt et al., 2021).

The research was approved by the Institutional Review Board of Pennsylvania State University (STUDY00009589) and the Kenya Medical Research Institute (KEMRI/RES/7/3/1) and followed the Declaration of Helsinki. Permission was also obtained from the Director of Health in the county government of Marsabit, Kenya and from community leaders. All participants provided written and verbal informed consent.

### Household Water Insecurity

Household water insecurity was measured using the 12-question household water insecurity experiences (HWISE) scale, which has been cross-culturally validated in low- and middle-income countries, including Kenya (Young et al., 2019). We worked with Daasanach research assistants to translate the survey into Daasanach. The HWISE items describe the frequency of different water-related experiences (e.g., inability to bathe, going to bed thirsty) that occurred over the past four weeks. Each item was scored from 0-3 for responses of never (0 times), rarely (1-2 times), sometimes (3-10 times), and often/always (11+ times), respectively, with the score range between 0 and 36. We categorized scores 0-11 as water secure, 12-23 as moderately water insecure, and 24-36 as severely water insecure. The HWISE scale was reliable in this context (Cronbach’s Alpha = 0.88).

### Psychosocial Stress

Psychosocial stress in the prior four weeks was measured among all household heads using the 4-item version of the validated Cohen Perceived Stress Scale (PSS-4). The PSS-4 has been used in multiple settings in conjunction with the HWISE scale, including Kenya (Young et al., 2019) and other water insecurity studies (Tallman, 2019). The PSS-4 asks individuals to recall the frequency in which they found their life over the past four weeks to be unpredictable, unmanageable, uncontrollable, uncertain, and/or overloaded (Cohen et al., 1983). Scores for each item ranged from 0-4 for responses of never, almost never (1-2 times), sometimes (3-10 times), fairly often (11-20 times), and very often (>20 times), respectively. Two positively phrased questions were reverse-coded, and scores for the items were summed for a total PSS-4 score range of 0-16 (Cohen et al., 1983).

We also measured stress for the prior 24 hours using the validated Daily Inventory of Stressful Events (DISE) following Almeida et al. (2002). The items asked about seven different stressful events (e.g., having an argument with anyone) since “this time yesterday.” Scores for each question were affirmations (0 or 1); summed DISE scores ranged from 0-7. Both stress scales PSS-4 and DISE were asked at the individual level.

### Water Borrowing and Water Lending

Households were asked during surveys, how frequently they borrowed water from people outside their household in the prior four weeks, following Rosinger et al., (2020). Water lending was assessed in the same manner. Responses for both questions were coded using the same options as the HWISE items, never (0 times), rarely (1-2 times), sometimes (3-10 times), and often/always (11+ times). For analytical purposes (i.e., cell sizes), we grouped the responses for never and rarely into one category.

When participants answered affirmatively to either borrowing or lending water, we further inquired whether the person to whom they lent water or from whom they borrowed water was family living nearby/neighbors, family not living nearby, neighbors (not family), friends (not neighbors), or other.

### Covariates

We adjusted for a number of covariates in our analyses due to their association with either water insecurity, water borrowing, or perceived stress.

Sex was included because studies have shown that males and females have differing experiences with water insecurity (Wutich, 2009; Brewis et al., 2019a). Among sub-Saharan African countries, adult women are the primary water collectors (Graham et al., 2016), as is the case among Daasanach. Hence, women may be more likely than men to experience stress resulting from water insecurity (Brewis et al., 2019a).

Studies also show that households experience and cope with water insecurity differently based on their wealth and socioeconomic status (Achore et al., 2020; Venkataramanan et al., 2020). To account for these differences three measures of socioeconomic status were used: livestock wealth, household income, and perceived standing in the community. To serve as an indicator of household wealth, we asked households about the number of each type of animal they owned and multiplied that number by their approximate value and summed the total as outlined in Rosinger and colleagues (2021). We further asked about and summed all income earned from any household members from any sources in the prior month. Finally, perceived socioeconomic status (SES) was measured using a MacArthur Scale of Subjective Social Status (Adler et al., 2008; Giatti et al., 2012). This pictorial tool allows an individual to select their status within the community as it related to wealth, education, and social status using 10 ladder rungs as a ranking (1 being the best off and 10 being the worst off).

Body mass index (BMI), an indicator of nutritional status, was calculated using participant weight to the nearest 0.1 kg and height to the nearest 0.1 cm. Weight was measured using a bio-impedance scale (Tanita BF-680W). Height was measured without shoes using a portable Seca standing stadiometer placed on a hard flat surface.

Age in years was self-reported and confirmed with an ID card when present. Since many Daasanach do not have a recorded date of birth, age was estimated based on birth around a historical event if necessary. The age of the female household head was used as a control in analyses conducted at the household level since Daasanach women are responsible for water.

Poor water quality is associated with higher levels of water insecurity and subsequent negative health outcomes (Bennyworth et al., 2016; Rosinger, 2018). Water salinity, one measure of poor water quality and a key concern among Daasanach (Rosinger et al., 2021), was measured using an YSI ProDSS Multi-Parameter Water Quality Meter and accompanying sensors. This meter provides a measure of the total concentration of dissolved salts in water. The recommended taste threshold for sodium is 200 mg/L (WHO, 2017).

We also constructed a perceived water quality variable from 2 survey questions about the number of times in the last 4 weeks anyone in the household consumed water that looked, tasted, or smelled bad and the number of times they worried the drinking water would cause sickness. The summed perceived water quality scores were calculated in a similar manner as the HWISE scale and ranged from 0-6.

Respondents were asked to report the amount of time a usual round-trip to fetch water including queue time, which sometimes may include time spent digging or re-digging a well, took them. Further, they reported the total number of trips the household took to collect water in the previous week.

Household size and composition were determined by asking how many children 7 years old and under, children 8-16 years, and adults age 17+ years lived in the household. Following a procedure similar to methods used for estimating adult male equivalents for comparing households overall caloric needs (Weisell & Dop, 2012), we used information on household members of each age group combined with European Food Safety Authority (EFSA) water recommendations (EFSA, 2010) to estimate average water needs. We used these values to create an adult equivalent household size for which we multiplied the number of 8- to 16-year old children by a corrective factor of 0.85 and the number of children <8 years old by a factor of 0.58. We summed these numbers with the household members >16 years old.

Finally, given traditional practices of migrating with livestock to meet water needs, respondents were asked the number of times they moved or traveled with their livestock over the past year. This number was used as an indicator of which households had higher mobility and practiced a nomadic lifestyle. Those with higher mobility are predicted to have lower water insecurity because of their ability to move to meet their water needs.

### Statistical Analysis

All analyses for this study were performed using Stata V.15.1 (College Station, TX). Our analytical sample included 133 households and 233 adults ≥18 years old with complete data for our variables of interest. For regression analyses, we used robust standard errors clustered by community of residence or household for household-level and individual-level models, respectively. For all models we adjusted for community fixed effects.

For our first aim, we examined how household and environmental characteristics were associated with the HWISE scale using a tobit regression since the outcome is censored at 0 and 36. Our model included the covariates for drinking water salinity, perceived water quality score, time spent fetching water, number of water fetching trips, household monthly income, livestock wealth, perceived SES ladder, household size adjusted for water needs, number of times the household moved, and the age of female household head. This analysis was conducted at the household level.

For our second aim, we dichotomized borrowing water and used logistic regression to estimate how HWISE score was associated with the odds of borrowing water never/rarely compared to those who do it more frequently (≥3times) at the household level. We controlled for the same covariates as in the prior model. We then used marginal standardization to estimate the predicted probability of borrowing water by HWISE score adjusting for the distribution of covariates (Muller et al., 2014).

For our third and final aim, we used tobit regression analysis to determine if water borrowing moderates the relationship between household water insecurity and psychosocial stress (PSS-4) at the individual level. We tested an interaction term between continuous HWISE score and level of water borrowing. Covariates included in the analysis were sex, livestock wealth, perceived SES, BMI, and individual age (Graham et al., 2016; Achore et al., 2020). Using marginal standardization as described above, we visualized the interaction between water borrowing and HWISE score on predicted PSS-4 scores based on the results of the model. Finally, as a robustness analysis, we re-estimated this analysis with the DISE daily stressor score in place of the PSS-4 score.

## Results

**Table 1** summarizes data for the demographics and covariates of the study households (n=133). Approximately 5% were water secure (HWISE scores 0-11), 61% were moderately water insecure (HWISE 12-23), and 34% were severely water insecure (HWISE ≥24) (**Figure 1**). The mean water insecurity score was 20.2 (±SD=6.7). The average water salinity of water sources used by households for drinking was 366 mg/L (±119) (**Table 1**). Approximately 97% of adult respondents spent more than one hour for a single trip to fetch water (including waiting time), while 77% spent two or more hours, for a mean of 121 (±45) minutes per trip (**Table 1**).

**Figure 1:**
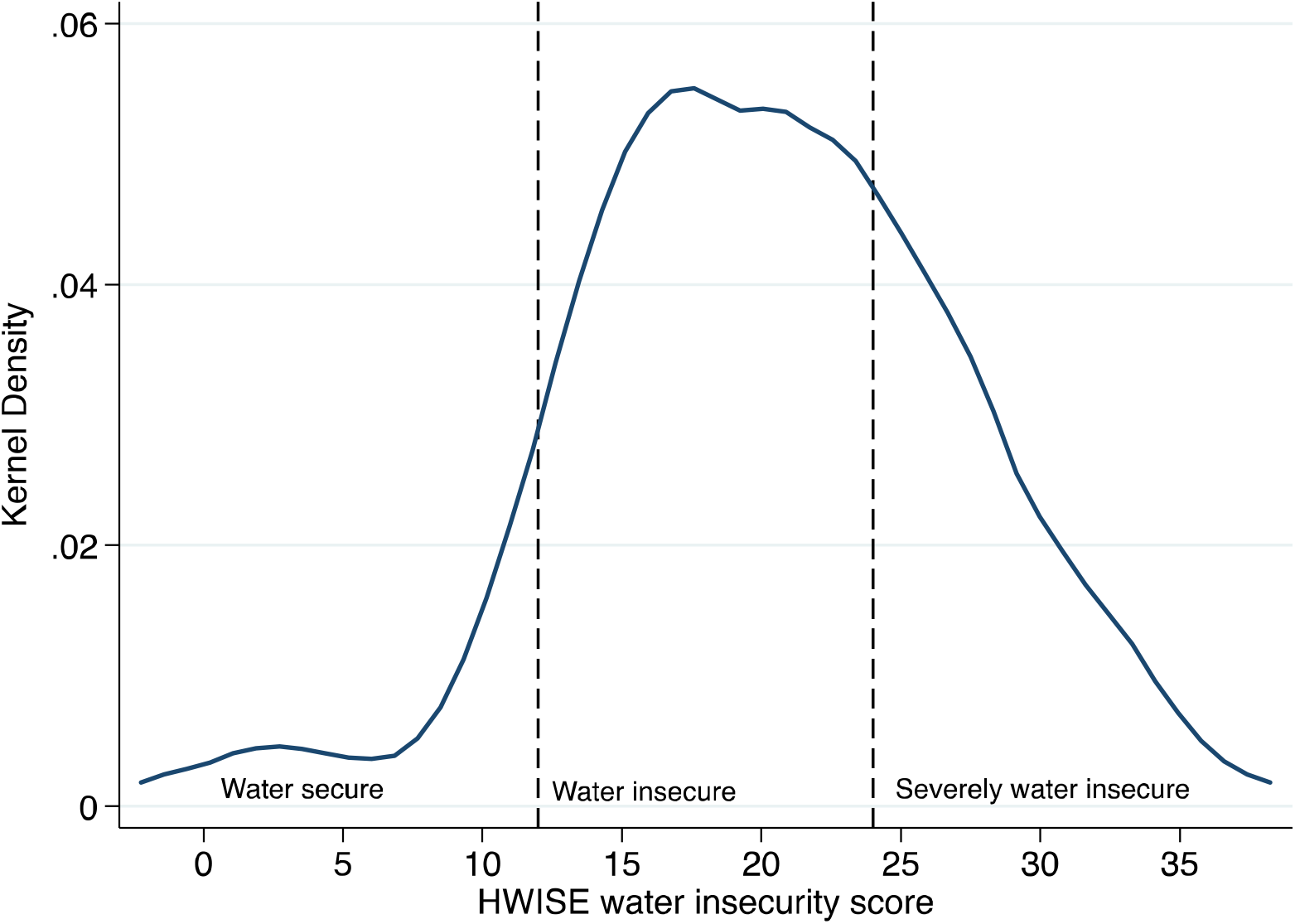
Kernel density of Water Insecurity HWISE score and categories of water insecurity (n=133)

**Table 1:** Descriptive Characteristics of Daasanach Households and Household Heads.

|  | Households (n=133) |  |
| --- | --- | --- |
|  | Mean or % | SD |
| HWISE score<sup>1</sup> (score range: 0-36) | 20.4 | 6.7 |
| Perceived water quality score<sup>2</sup> (score range: 0-6) | 2.3 | 1.6 |
| Borrowed water: never/rarely (0-2 times) | 25.6% |  |
| Sometimes (3-10 times) | 45.9% |  |
| Often/always (11+ times) | 28.6% |  |
| Lent water: never/rarely (0-2 times) | 14.3% |  |
| Sometimes (3-10 times) | 48.9% |  |
| Often/always (11+ times) | 36.8% |  |
| Salinity (mg/L) | 366 | 119 |
| Time spent fetching water per trip (minutes) | 121 | 44.6 |
| Weekly water trips | 15.5 | 6.9 |
| Times moved in last year | 4.2 | 6.2 |
| Household size modified by water need | 5.9 | 2.2 |
| Livestock wealth (USD) | 2002.5 | 5367 |
| HH monthly income (Kenyan Shillings) | 3400 | 9798 |
| Perceived SES ladder (1-10) | 7.5 | 2.3 |
|  | Adults (n=233) |  |
|  | Mean or % | SD |
| Age (years) | 40.2 | 15.0 |
| PSS-4 score (score range: 0-16) | 7.9 | 2.2 |
| Daily stress DISE score (score range: 0-7) | 0.92 | 1.1 |
| BMI (kg/m<sup>2</sup>) | 18.3 | 3.1 |
| Male (%) | 45.9% |  |

<sup>1</sup>HWISE = Household Water Insecurity Experiences
HH: household; SES: Socio-economic status

It was common among respondents to borrow and lend water; 74.4% of respondents borrowed water three or more times in the previous four weeks, while 85.7% lent water ≥3 times (**Figure 2a-b**). Those who borrowed water were most likely to borrow from their neighbors (86%), while those who lent water were also likely to give water to their neighbors (75%) (**Figure 2c-d**). The majority of respondents both borrowed and lent water (63%).

**Figure 2:**
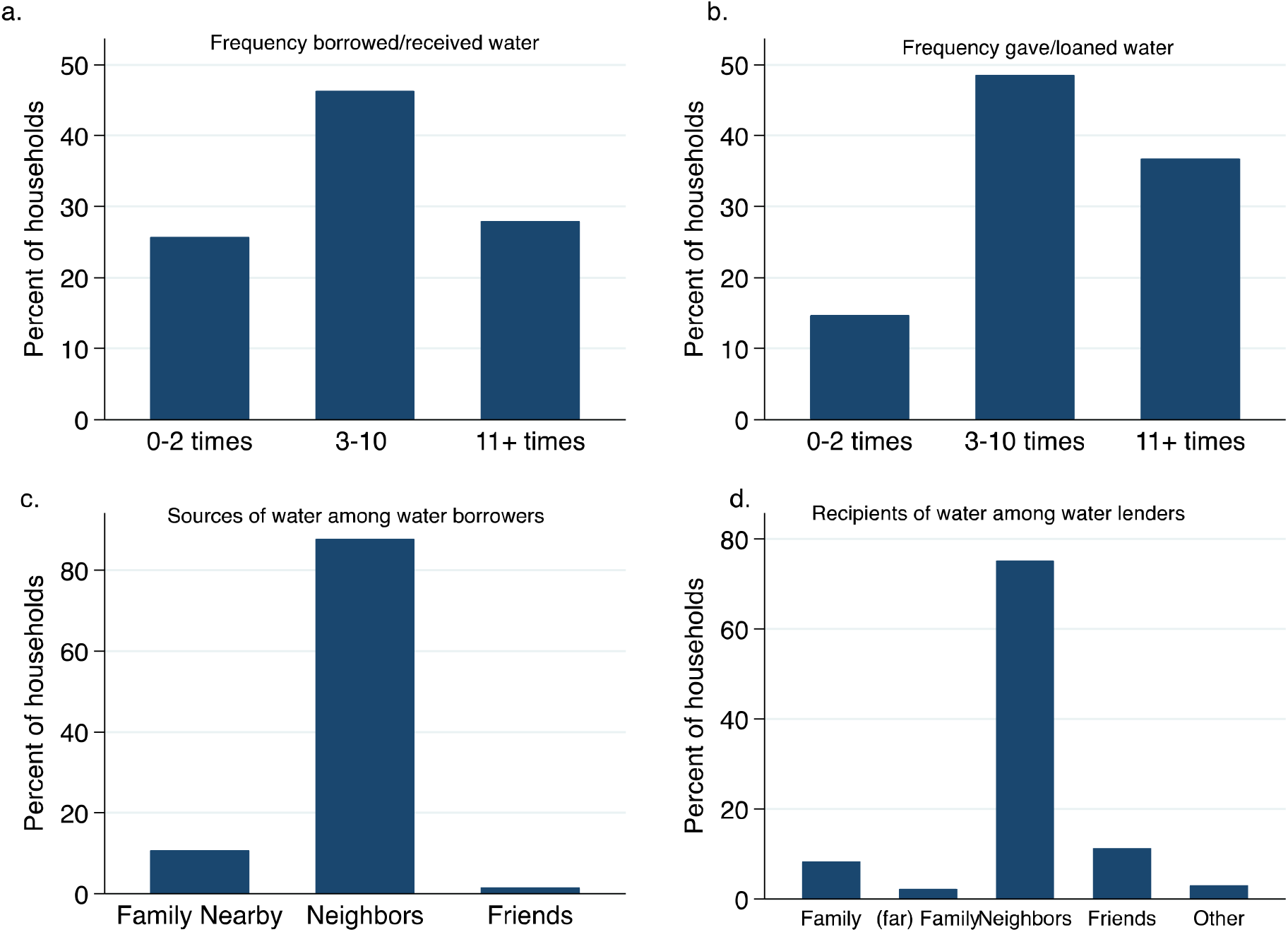
Water Borrowing among Daasanach (a) frequency of water borrowing, (b) frequency of water lending/giving, (c) reported givers/sources of water among those who borrowed water, and (d) reported recipients of water among water lenders.

### Aim 1: Predictors of water insecurity

The results of the tobit regression indicate that several household and environmental factors were associated with HWISE score (**Table 2)**. Objective water quality as measured by water salinity was found to be significantly associated with water insecurity, each 100mg/L increase in salinity was associated with 1.58-points (SE=0.40, P<0.001) greater HWISE score. Perceived water quality was also strongly associated with water insecurity; each point higher was associated with 2.03-points (SE=0.55, P<0.001) higher HWISE score. The accessibility of water was another significant predictor. Both time spent fetching water (β =0.30 per 10 minutes, SE=0.11, P=0.007) and number of weekly trips (β =0.56 per 3 trips, SE=0.27, P=0.04) were significantly associated with HWISE scores. The only other predictor significantly associated with water insecurity was household size adjusted for water needs as each additional adult equivalent was associated with 0.64-points (SE=0.21, P=0.003) higher HWISE score. SES variables, monthly income, SES Ladder, and livestock wealth were not associated with water insecurity.

**Table 2:**
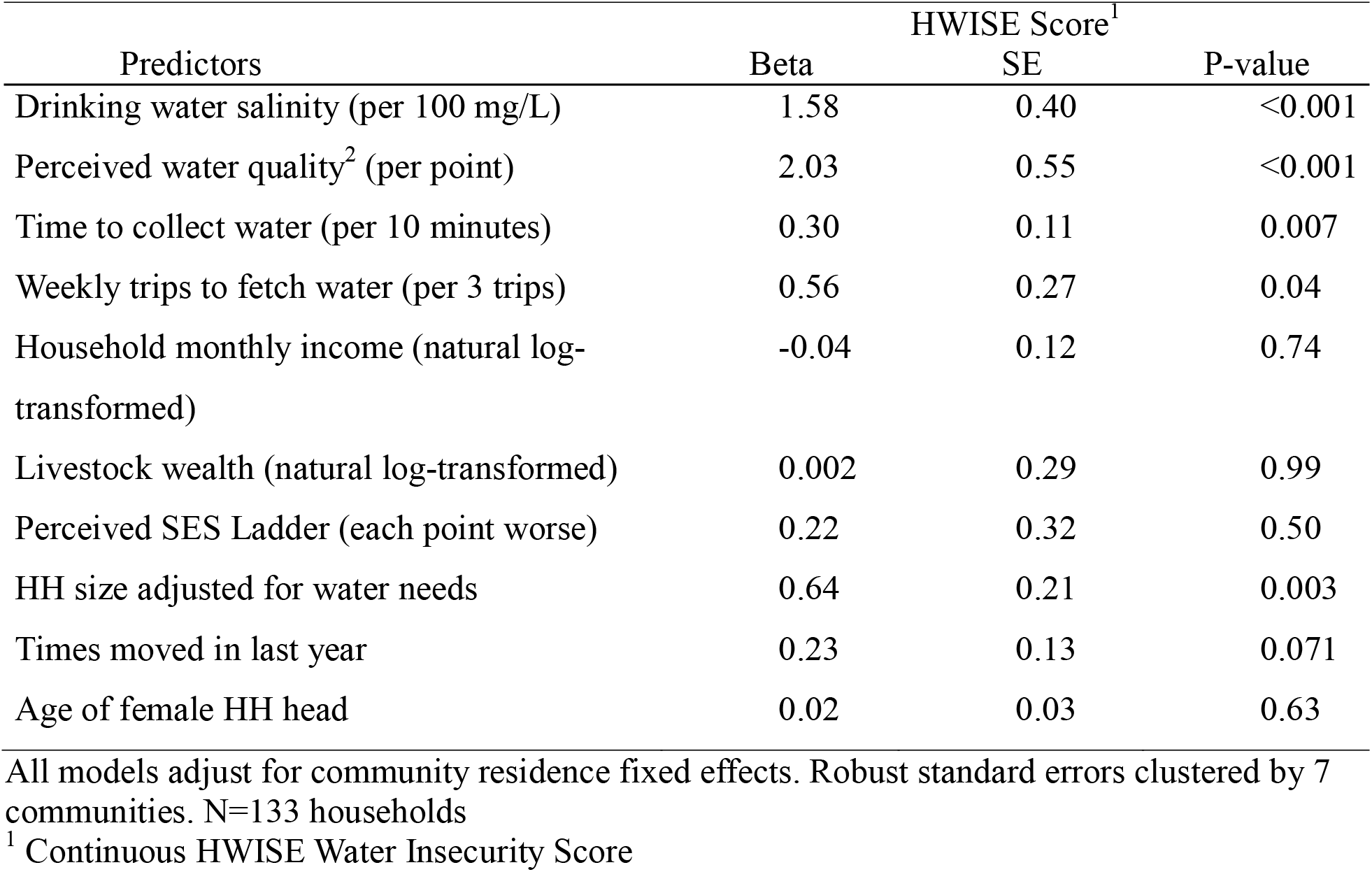
Tobit regression examining predictors of water insecurity among Daasanach.

### Aim 2: Water insecurity and water borrowing

Results of the logistic regression demonstrate that HWISE score was strongly associated with the odds of borrowing water 3 or more times in the prior 4 weeks. Each additional point on the HWISE scale was associated with 16% higher odds of water borrowing (OR=1.16; 95% CI=1.04-1.29; P=0.006) adjusted for other household and environmental covariates (**Table 3**). Lower SES was associated with relying on borrowing water more frequently. Each point worse on the ladder was associated with 29% higher odds (OR=1.29; 95% CI=1.02-1.64; P=0.03) of borrowing water more frequently.

**Table 3:** Logistic regression examining predictors of water borrowing among Daasanach households.

| VARIABLES | Borrowed Water |  |  |
| --- | --- | --- | --- |
|  | Odds Ratio | 95% CI | P-value |
| HWISE Score (per point) | 1.16 | 1.04-1.29 | 0.006 |
| Drinking water salinity (per 100 mg/L) | 0.99 | 0.09-11.2 | 0.99 |
| Perceived water quality | 1.47 | 0.80-2.71 | 0.22 |
| Time to collect water (per 10 minutes) | 1.08 | 0.95-1.24 | 0.22 |
| Weekly trips to fetch water (per 3 trips) | 1.02 | 0.93-1.11 | 0.72 |
| HH monthly income (natural log-transformed) | 0.91 | 0.77-1.06 | 0.23 |
| Livestock Wealth (natural log-transformed) | 0.92 | 0.74-1.15 | 0.45 |
| Perceived SES Ladder<br>(each point worse) | 1.29 | 1.02-1.64 | 0.03 |
| HH size adjusted for water needs of age groups | 1.02 | 0.88 -1.18 | 0.81 |
| Age of female HH head | 0.97 | 0.93-1.01 | 0.10 |

n=133 households
Model adjusted for community fixed effects. Standard errors clustered on seven communities.
Outcome borrowed water is dichotomized as $\geq 3$ times compared to 0-2 times.

**Table 4:**
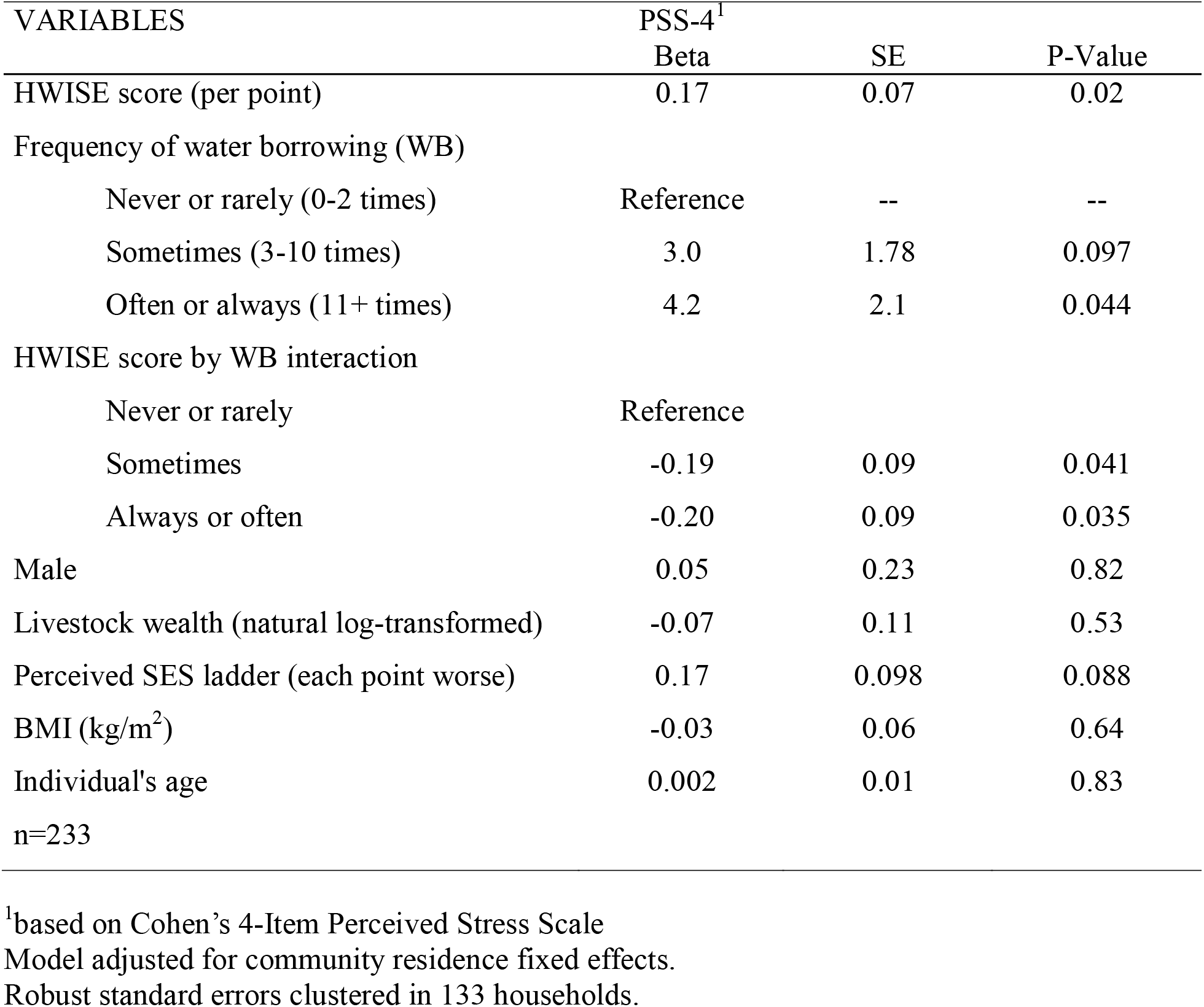
Tobit regression examining the association between water insecurity and psychosocial stress testing water borrowing as a possible moderator.

Water borrowing was tightly connected to water insecurity score. The predicted probability of borrowing water increased from 38.7% at an HWISE score of 6, to 56.6% at 12, and to 84.4% at an HWISE score of 24 (**Figure 3**).

**Figure 3.**
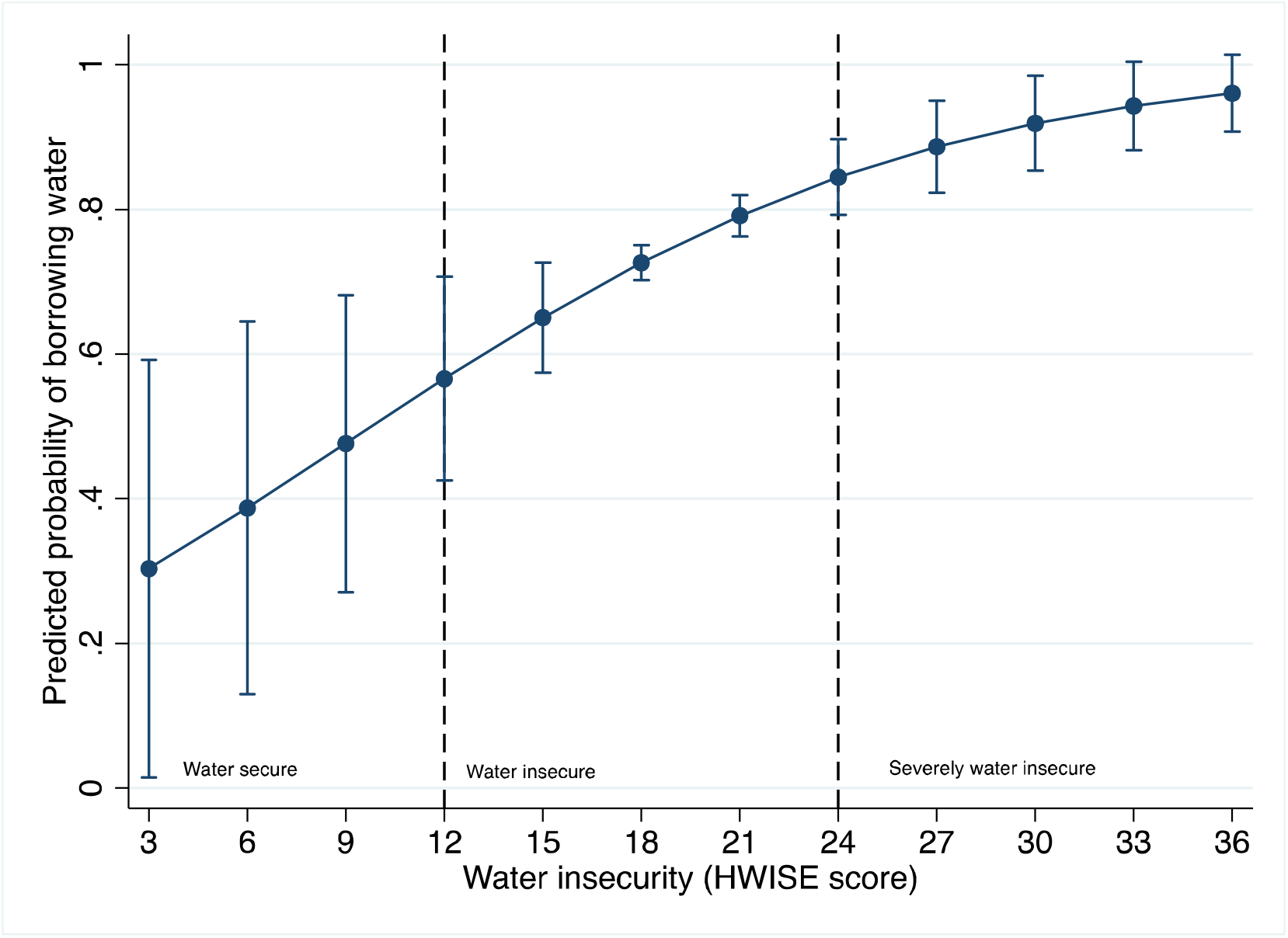
Predicted probabilities and 95% confidence intervals of borrowing water 3 or more times in prior 4 weeks by water insecurity score. Notes: Adjusted for the range of covariates presented in the model presented in Table 3.

During follow-up discussions with communities about water borrowing, there was consensus that they often borrow water during the dry season because they can become tired of searching for water in the dry riverbeds. They are however, expected to repay when they obtain their water. Yet, others indicated that they share water with neighbors during the wet season, when it is more plentiful with no expectation for payback, but not always during the dry season. Thus, water borrowing may relate to seasonality, water availability, along with the expectation of return.

### Aim 3: Water borrowing, water insecurity, and PSS-4

In bivariate analyses, HWISE score was significantly correlated with PSS-4 (r=0.18, P=0.006) (**Figure 4**). In the multiple tobit regression analysis, the main terms of HWISE score (β =0.17, SE=0.07, P=0.02) and borrowing water often/always (β =4.2, SE=2.1, P=0.044) were both associated with higher PSS-4 scores. Further, there was a significant interaction between frequency of water borrowing and HWISE score. Compared to those who never or rarely borrowed water, those who borrowed water sometimes and often/always had 0.19 (SE=0.09, P=0.041) and 0.20 (SE=0.09, P=0.035) lower PSS-4 scores, for each point higher on the HWISE scale.

**Figure 4:**
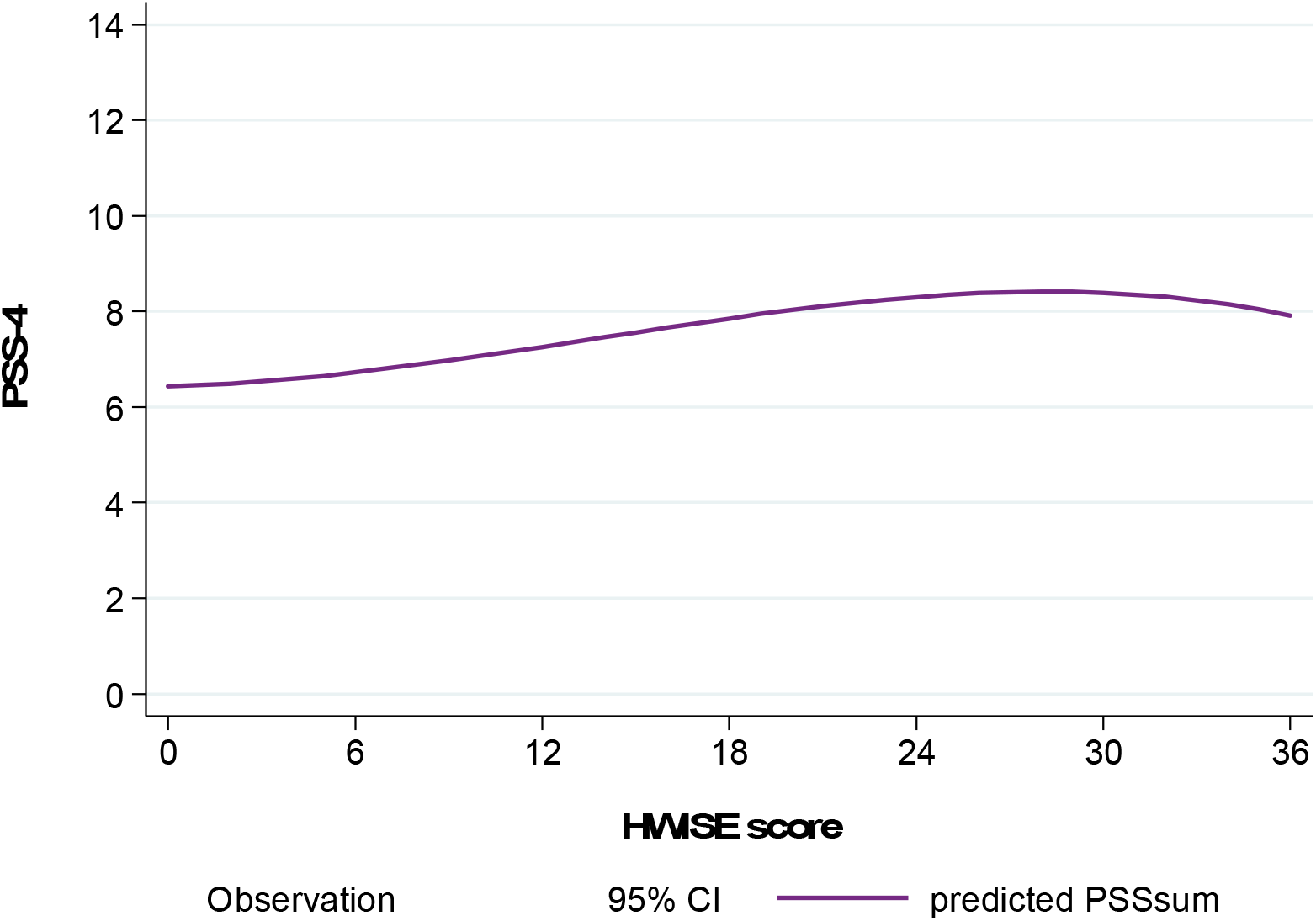
Scatterplot of HWISE scores on perceived stress scores and 95% confidence interval. PSS-4: Cohen’s 4-Item Perceived Stress Scale

This significant moderation effect demonstrates significantly different slopes between the levels or categories of water borrowing frequency on PSS-4 as water insecurity increased (**Figure 5**). It indicates that those who borrowed water never or rarely saw a linear increase in PSS as HWISE score increased, whereas for those engaged in higher levels of water borrowing, the relationship was slightly negative.

**Figure 5:**
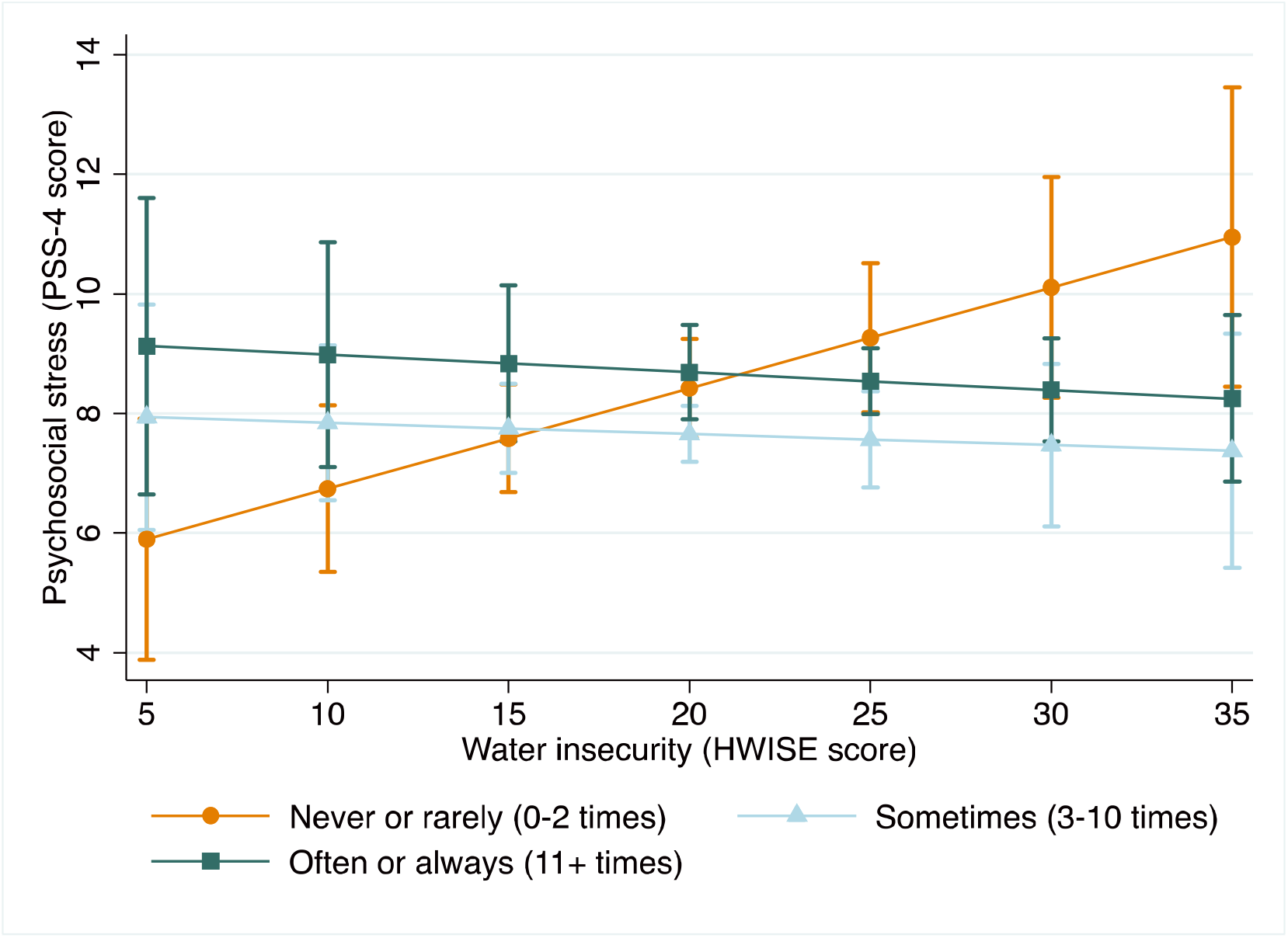
Predicted psychosocial stress by water borrowing in last four weeks and HH water insecurity score.

### Robustness analysis

Re-estimating the analysis relationship between water insecurity, stress, and water borrowing using the DISE inventory of daily stressors score in place of the PSS-4 score, we find consistent results despite the change in timescales from the previous month to the previous day (**Supplemental Table 1**). Those who never or rarely borrowed water had a linear increase in daily stressors as HWISE score increased, whereas those who borrowed water sometimes and often/always had a negative association between HWISE score and DISE score (**Supplemental Figure 1**).

## Discussion

This study aimed to 1) quantify water insecurity among Daasanach pastoralists living in an extreme hot-arid environment and examine how environmental and household factors are associated with household water insecurity, 2) examine how water insecurity is associated with water borrowing, and 3) evaluate whether water borrowing acts as a coping mechanism to moderate the association between household water insecurity and individual psychosocial stress.

### Water insecurity among Daasanach

Water insecurity was nearly ubiquitous among Daasanach households as 95% of households were classified as moderately or severely water insecure according to the HWISE scale. It is not clear how many other populations experience similarly high prevalence of water insecurity, though at least two other sites (Punjab, Pakistan and Cartagena, Colombia) reported mean HWISE scores on par with our sample (Stoler et al, under review). Other studies among underserved pastoral populations in Sub-Saharan Africa including Ethiopia (Stevenson et al., 2012) and Uganda (Pearson et al., 2015), and Northern Kenya (Balfour et al., 2020) have similarly documented high levels of water scarcity and water insecurity.

Among Daasanach households, indicators of water quality, access, and quantity were all significantly associated with experiences of water insecurity. Specifically, indicators of water salinity and perceived water quality were associated with higher HWISE scores. This finding is consistent with a study conducted in Bangladesh, which found that slightly saline groundwater and water treatment practiced by approximately half of the respondents was associated with elevated water insecurity (Benneyworth et al., 2016). Among Daasanach, the average drinking water salinity level was 360 mg/L, which is above the recommended taste threshold for sodium of 200 mg/L. Further, our salinity findings map onto local concerns expressed during fieldwork about the salinity of the groundwater (Rosinger et al., 2021). While there are currently no health guideline values for sodium (WHO, 2017), prolonged consumption of saline water has been linked with negative health outcomes such as elevated systolic and diastolic blood pressure, increased risk of hypertension, reproductive concerns for women, and altered cognitive performance (Rosinger et al., 2021).

Our measures of water accessibility, time spent fetching water and number of water fetching trips per week, were both significantly associated with water insecurity score. Other household factors, like household size, were also associated with water insecurity as a marker of increasing water need. In contrast with our expectations, SES variables were not associated with HWISE score. Yet, prior work indicates that not only does the time required to fetch water directly contribute to water insecurity, it can also indirectly contribute to it by reinforcing economic barriers that prevent household from sustainably addressing water insecurity concerns (Achore et al., 2020).

### Water borrowing as a response to water insecurity

We found that ∼75% of Daasanach households borrowed water at least 3 times in the prior month and that water insecurity was strongly associated with water borrowing frequency. The fact that the majority of households that borrowed water also lent water, and that both of these occurred primarily among neighbors indicates that reciprocity may be an important consideration in the water-borrowing network. This is consistent with Daasanach culture of borrowing and sharing goods and food to benefit the community at large (Wright, 2019).

Historical and ethnographic records from other populations, including pastoralist records also suggest that water sharing is a common strategy for meeting water needs in times of extreme water shortages globally (Wutich et al., 2018). In the largest study documenting this, water borrowing ranged from 11-85% across 21 sites with known water problems (Rosinger et al., 2020). The relatively high prevalence of water borrowing across diverse, water-stressed environments as a strategy to mitigate water insecurity and water system failures highlights the need to understand how these practices affect the social and cultural norms, which influence water acquisition, distribution, and prioritization (Rosinger et al., 2020). Other studies indicate that reciprocity of water between wealthy and poor households is critical for maintaining social capital between these groups (Pearson et al., 2015). This may also be a mechanisms through which social power relations are maintained (Wutich et al., 2018; Pearson et al., 2015).

### Water borrowing as a moderator of stress

Water insecurity was significantly associated with psychosocial stress in our study, a finding consistent with our original hypothesis and similar to findings from other studies (Collins et al., 2019; Brewis et al, 2019a; Stoler et al., 2019; Wutich, 2009; Stevenson et al., 2012). In many contexts household water managers often experience psychosocial stress as a result of the negotiations, such as borrowing, purchasing, or rationing water, in which they must engage to ensure sufficient water (Wutich & Ragsdale, 2008; Brewis, 2019a; Stevenson et al., 2012). Marginalized households with low socioeconomic status often rely on less costly but more time-consuming coping mechanisms for water insecurity, which may contribute to even greater stress among those households (Achore et al., 2020; Venkataramanan et al., 2020). We found that lower perceived standing was associated with higher odds of borrowing water. Daasanach frequently noted during interviews that they would borrow water from neighbors when they did not have enough time to fetch their own. One woman told us that if she lacked water in the late afternoon, she would often ask a neighbor for water to cook dinner with and then repay that water in the future.

Among Daasanach, the frequency of water borrowing moderated the relationship between water insecurity and psychosocial stress. For households that never or rarely borrowed, higher water insecurity was associated with significantly higher perceived stress; while for those engaged in water borrowing more frequently, greater water insecurity was slightly negatively associated with perceived stress (**Figure 5**). While at low levels of water insecurity, those who never or rarely borrowed water had lower stress than individuals who relied on water borrowing, at higher levels of water insecurity they had higher stress than those who borrowed water. These findings are consistent with theory that suggests water sharing can lower psychosocial stress by providing safety nets for those households who experience water insecurity (Stoler et al., 2019; Wutich et al., 2018).

Our results further suggest that water borrowing is an effective coping mechanism for those who are included in the water borrowing network, but people who do not borrow water, for whatever reason, may suffer more stress than those who borrow water. This finding echoes the theoretical underpinnings of water borrowing outlined in Wutich et al. (2018) which suggests that while water sharing may reduce levels of psychosocial stress for those included in the network, those who are excluded may experience increased stress. Our results are also consistent with recent research from Ethiopia, which find that level of participation in informal water sharing networks is critical to understand in conjunction with unfairness for psychological distress (Brewis et al., 2021). As a result, water borrowing as a coping mechanism may be dependent on a household’s relationship with neighboring households and their inclusion in the network (Achore et al., 2020). Future work should untangle what determines networks and why certain households are or are not in-network and how distance to the main market town affect these relationships.

### Limitations

Limitations include that the survey was cross-sectional, such that results should be viewed as associations and not causal, though results were substantiated with follow-up focus group discussions. Data for the survey were collected during the early dry season but before water insecurity is arguably at its worst (around September). The water insecurity experiences captured during June and July may not be representative of the entire year because it does not capture seasonal variations and migration with livestock.

Second, while our measure of psychosocial stress (PSS-4) has been validated in different populations and we consulted with local informants about the appropriate translation and interpretation of these questions, it is possible that we were not capturing all the domains of psychosocial stress and mental health distress that members of the Daasanach community experience as a result of water insecurity. However, our results were substantiated by the DISE daily stress inventory which is an instrument that assesses actual stressor events rather than appraised subjective stress from the PSS.

Finally, we do not know if people who did not borrow water chose not to do so or were excluded from doing so. This is important because the effect on stress could differ depending on whether they feel resentful about having to borrow water, or about being excluded from the borrowing network. Future work should examine reasons for borrowing, its relationships with water security, and complement this work with qualitative data to ensure that nuances are understood.

## Conclusion

In conclusion, this study demonstrated that Daasanach pastoralists experience high levels of household water insecurity during the early dry season and that water insecurity predicts higher frequency of water borrowing. Further, water borrowing frequency has important implications for Daasanach psychosocial stress experienced in association with water insecurity. We found that the positive relationship between water insecurity and psychosocial stress was not observed among those who borrowed water, suggesting that borrowing water may serve to reduce the psychosocial burden of water insecurity. These findings are valuable because they contribute to a small but growing body of literature that seeks to quantify household level water insecurity experiences among pastoralist and semi-nomadic groups in the arid regions of eastern Africa (Pearson et al., 2015; Balfour et al., 2020). Additionally, these findings are among the first to explore the interaction between household level water insecurity and water borrowing as a coping mechanism to mitigate psychosocial stress. Future studies investigating water insecurity and psychosocial stress should consider water borrowing as a possible moderator in other populations. Water borrowing may serve as a powerful informal system to blunt psychosocial stress from water insecurity for those included in water borrowing networks.

## Data Availability

All data produced in the present study are available upon reasonable request to the authors

## Acknowledgements

We thank Luke Lomeiku, Samuel Esho, and Joshua Koribok, and the community health volunteers that helped with data collection. We thank Purity Kiura, The Koobi Fora Field School, and The National Museums of Kenya for facilitation with the project. We thank the Illeret Health clinic, The Illeret Ward administrator Mr. Koriye Koriye, and all of the Daasanach communities and participants. Finally, we thank research assistants (Celine LaTona, Alysha Kelyman, Kaitlyn Barnhart, Jason John) of the Water, Health, and Nutrition Lab for their help in data cleaning.

---

**Supplemental Figure 1:**
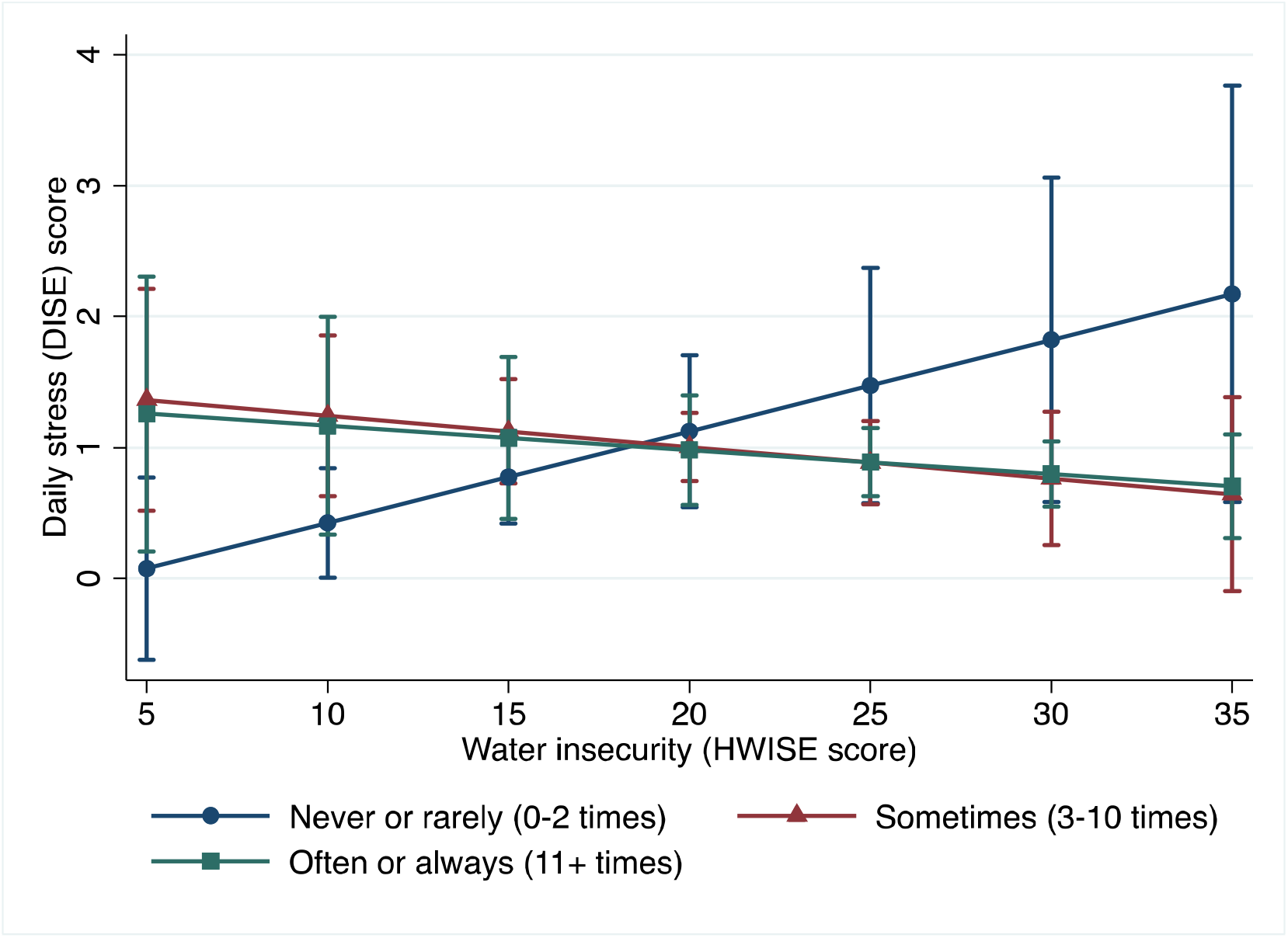
Predicted daily stress by water borrowing level and HH water insecurity score.

**Supplemental Table 1:** Regression examining the association between water insecurity and daily stress score with water borrowing as a possible moderator.

| VARIABLES | DISE<sup>1</sup><br>Beta | SE | P-Value |
| --- | --- | --- | --- |
| HWISE score (per point) | 0.07 | 0.036 | 0.057 |
| Frequency of water borrowing (WB) |  |  |  |
| Never or rarely (0-2 times) | Reference | -- | -- |
| Sometimes (3-10 times) | 1.76 | 0.74 | 0.019 |
| Often or always (11+ times) | 1.62 | 0.87 | 0.065 |
| HWISE score by WB interaction |  |  |  |
| Never/rarely | Reference |  |  |
| Sometimes | -0.09 | 0.04 | 0.03 |
| Always or often | -0.09 | 0.045 | 0.055 |
| Male | 0.18 | 0.14 | 0.21 |
| Perceived SES ladder (each point worse) | -0.05 | 0.05 | 0.37 |
| Livestock wealth (log-transformed) | -0.05 | 0.04 | 0.23 |
| BMI (kg/m<sup>2</sup>) | 0.05 | 0.024 | 0.064 |
| Individual's age | -0.0005 | 0.01 | 0.93 |
| n=233 |  |  |  |

<sup>1</sup> Daily stress score based on the 7 Item DISE Scale
Results are from a tobit regression model that regresses psychosocial stress score with the predictors listed above. Adjusted for community residence fixed effects. Robust standard errors clustered in 133 households.

